# Deceptive Bias Measurement in Deep Learning: Assessing Shortcut Reliance in TCGA Cancer Models

**DOI:** 10.64898/2025.12.11.25342109

**Authors:** Farnaz Kheiri, Shahryar Rahnamayan, Masoud Makrehchi

## Abstract

Bias in machine learning is a persistent challenge because it can create unfair outcomes, limit generalization, and reduce trust in real-world applications. A key source of this problem is shortcut learning, where models exploit signals linked to sensitive attributes, such as data source or collection site, instead of relying on task, relevant features. To tackle this, we propose the Deceptive Signal metric, a novel quantitative measure designed to assess the extent of a model’s reliance on hidden shortcuts during the learning process. This metric is derived via the Deceptive Bias Detection pipeline, which isolates shortcut dependence by contrasting model behavior under two controlled conditions: (1) Full Exclusion, where a sensitive subgroup is completely removed from training; and (2) Partial Exclusion, where the model has limited access to specific classes within the subgroup. By calculating the behavioral shift between these settings, the Deceptive Signal metric provides a concrete value representing the model’s proneness to learning task-irrelevant patterns. In experiments with the TCGA histopathology dataset, our metric successfully quantified strong dependencies on center-specific artifacts in models trained for cancer classification.

**Author summary:** Deep learning models are becoming powerful tools in healthcare, but they often suffer from a critical vulnerability: they can get the right answer for the wrong reason. In medical imaging, an AI might correctly identify a tumor not by analyzing the tissue, but by recognizing irrelevant digital markers unique to the specific hospital or scanner that produced the image. This phenomenon, known as shortcut learning, makes AI systems appear accurate at first glance while remaining unreliable for real-world patient care.

To solve this, our research moves beyond simple accuracy checks and introduces a specific quantitative metric for shortcut learning. We developed a testing framework that forces the model into controlled training scenarios, deliberately withholding specific “shortcut” information to see how the model reacts. By mathematically comparing the model’s behavior across these scenarios, we calculate a precise score that indicates the magnitude of the model’s dependence on irrelevant patterns. This metric allows to put a concrete number on a model’s trustworthiness and ensuring that medical decisions are driven by biology, not background noise.

## 1 Introduction

In recent years, the increasing integration of intelligent systems into various aspects of daily life has underscored the critical need for developing models that are both fair and unbiased. Bias in machine learning causes serious risks; it can produce discriminatory outcomes, compromise the model’s generalization to unseen data, and raise significant ethical and societal concerns [1–3]. To ensure fairness, models must rely on task-relevant features rather than exploit shortcuts associated with sensitive factors such as gender, or the origin of the data (e.g., specific data collection sites or institutions). As the foundation of any predictive system, training data plays an important role in shaping model outputs. Therefore, the presence of easy-to-learn but task-irrelevant patterns associated with sensitive attributes may severely undermine the model’s trustworthiness, particularly when it is deployed in real-world settings or evaluated on external samples that contain sensitive attributes not represented during training [4, 5].

One critical step during the training process is to assess whether the model is truly learning from task-relevant patterns or if its predictions are influenced by task-irrelevant shortcuts. These shortcuts often stem from spurious correlations or hidden signatures associated with sensitive attributes, such as demographic features or dataset origins [6, 7]. Evaluating how the model’s behavior changes in the presence or absence of these sensitive attributes is essential for identifying potential sources of bias. Such investigations help ensure that the model’s decision-making process remains aligned with the intended task rather than being changed by irrelevant or discriminatory patterns embedded in the training data [8, 9]. A notable example of this issue arises in medical imaging datasets [10], such as TCGA, where samples are collected from multiple data centers. These centers may differ in scanning devices, staining protocols, or other characteristics, unintentionally embedding identifiable patterns into the images. As a result, models may learn to associate such center-specific artifacts with the target labels (e.g., cancer type). As a result, it may lead to biased predictions that do not generalize well out of the training distribution [11, 12].

While many existing bias investigation methods focus on disparities in prediction outcomes across different sensitive groups, such as positive prediction rate [13]; they often overlook the underlying learning behavior during training. These approaches tend to assess bias after the model is trained, by measuring how its predictions vary across subgroups [14]. However, such outcome-based evaluations may fail to detect whether the model has relied on task-irrelevant features embedded within specific subgroups. To truly ensure fairness and generalization, it is crucial to go beyond surface-level performance metrics and analyze the training process itself by examining whether the model is learning representations that generalize to the task or simply memorizing subgroup-specific artifacts.

To contextualize the challenges of fairness and generalization, we review prior studies on bias detection and quantification. Here, we examine how recent research has attempted to assess these biases. We categorize existing methodologies into two primary streams: techniques that investigate hidden biases and spurious correlations within model representations, and outcome-based methods that evaluate fairness through disparities in predictive performance across sensitive subgroups.

### 1.1 Shortcut Learning and Hidden Bias in Representations

Here, we focus on studies that show how deep models may learn spurious correlations or shortcuts (e.g., background patterns, color artifacts, dataset source patterns) and employ these signatures to fulfill the target task [15]. Geirhos et al. [5] discuss that the models trained on the modified version of the original ImageNet dataset where textures were deliberately removed while object shapes were preserved performed significantly worse compared to models trained on original ImageNet. This reveals that the models relied heavily on texture-based cues during training.

Saliency-based methods are a popular approach widely used in prior studies to identify the regions or features within an input that most influence a model’s predictions. Techniques such as Grad-CAM [16–18] and saliency maps [19–21] aim to enhance the interpretability of model outputs by presenting which parts of the input data have the greatest impact on the model’s decisions during training or inference. The studies [22–28] applied saliency map techniques to medical images to investigate whether diagnostic decisions were based on task-relevant features or influenced by spurious, non-diagnostic patterns. The achieved results reveal that in many cases, deep models tend to focus on artifacts such as image borders, text annotations, or scanner-specific features, rather than the actual pathological regions.

Another widely used methodology is based on counterfactual scenarios [29], which aim to assess the fairness of model outputs by examining their sensitivity to changes in sensitive attributes. These methods involve modifying input features, typically by altering or removing sensitive variables such as gender or race [30–32], while keeping the target class constant. If the model’s prediction changes significantly after such a modification, it suggests that the model may be relying on task-irrelevant or sensitive features, rather than learning from the core features relevant to the task.

Autoencoders can help uncover shortcuts by learning a compact version of an image and reconstructing images to see what information and patterns are truly learned by the model. If the reconstruction removes irrelevant details (like scanner artifacts) and the main model’s accuracy drops, it suggests the model was relying on those irrelevant details rather than the real features [17, 33, 34].

### 1.2 Outcome-Based Fairness Evaluation

Another line of research in this section focuses on evaluating the impact of sensitive attributes on model outcomes using fairness metrics, typically by measuring disparities in predictive performance across different sensitive subgroups. These outcome-based fairness metrics operate under the assumption that a fair model should yield comparable performance or decision rates across all groups, regardless of which sensitive attributes they belong to. The method proposed in [35] applies the definition of group fairness metrics [14] to quantify the effect of encoded shortcuts on model outcomes. They measure disparities using metrics such as True Positive Rate or Equal Opportunity metric [36] across continuous sensitive attributes. The study in [37] employs Worst-Group Accuracy (WGA) to assess shortcut reliance by defining groups as combinations of labels and sensitive attributes. Lower WGA in fairer models suggests that removing shortcut signals can reduce performance on disadvantaged subgroups. Another research study [38] applied Equalized Odds metric to assign weights to balance class distributions within each demographic group to ensure comparable true positive and false positive rates. The research conducted in [27, 39] addresses the shortcut learning issue by reducing disparities in metrics such as Area Under the Curve, Binary Cross-Entropy, Expected Calibration Error, and Precision through a model and task-agnostic image augmentation strategy.

In this work, we address this gap by proposing a novel measurement metric designed to quantify the extent to which a model relies on task-irrelevant patterns associated with sensitive attributes. To derive this metric, we introduce the *Deceptive Pipeline*, a structured methodology that manipulates the training data to examine how these patterns influence model decisions and lead to biased behavior. We validate our approach using the TCGA dataset, which comprises samples collected from various medical centers. Prior studies [8, 12] have demonstrated that deep models trained on TCGA often capture data center-specific artifacts and leverage them as hidden shortcuts for cancer classification. Extending these findings, we analyze the training phase itself to assess the reliability of the learning procedure, determining whether the model is truly learning cancer-related characteristics or is driven by data center-specific signatures.

The Deceptive Pipeline facilitates this measurement by altering the presence or absence of specific sensitive attribute combinations via multiple training setups. This allows us to test whether the model learns task-relevant (cancerous) features or simply relies on task-irrelevant patterns tied to data center origin. By observing shifts in misclassification patterns across these controlled settings, we gain a clearer understanding of how bias emerges even before the evaluation phase. Consequently, the core contribution of this study is a quantitative metric that accurately reflects the model’s prone-ness to learning these task-irrelevant shortcuts.

Our proposed methodology consists of three structured phases to calculate this metric. **Phase 1 (Full Shortcut Exclusion)** involves training the model without any samples from a specific sensitive subgroup to assess its performance without exposure to subgroup-specific features. **Phase 2 (Partial Shortcut Exclusion)** selectively includes samples from only one class within the sensitive subgroup, allowing us to observe how partial exposure impacts classification behavior. Finally, **Phase 3 (Deceptive Signal Measurement)** compares the misclassification patterns between the full and partial exclusion settings to compute the proposed metric, characterizing the magnitude of the model’s dependence on task-irrelevant features.

## 2 Materials and methods

Advanced machine learning models, including deep neural networks, often rely on unintended correlations or shortcuts present in the input data to complete assigned tasks. These shortcuts may be associated with sensitive attributes, such as gender, or irrelevant patterns, such as unique signatures specific to certain data sources that are unrelated to the task’s actual objective. When models depend on such attributes, the outcomes become unreliable. This unreliability becomes more evident when testing input samples that lack these attributes are fed into the model, then it can lead to incorrect performance.

To clarify this concept with a real-world example, imagine a math teacher designing exam questions for students. These questions are nearly identical to those solved during tutorial sessions, with only minor numerical modifications. In such a case, exam results would primarily reflect how well students have memorized specific solutions, rather than their true understanding of the underlying concepts. The Deceptive Bias Detection method is designed to reveal such reliance by systematically modifying the models’ input to remove access to the sensitive signal and then reintroducing it during testing or inference to measure how model’s behavior is shifted.

### 2.1 Methodological Principle

The core hypothesis behind our Deceptive Bias detection pipeline is that if a model relies on hidden patterns or sensitive attributes as shortcuts to generate its output, its behavior will change significantly when those attributes are removed during training and then reintroduced during evaluation. Such a behavioral shift indicates that the model has encoded a dependency on those attributes. Depending on the case study we are working with, this hypothesis can be adapted to target specific types of bias, such as gender, source-specific artifacts, or domain-related features, by manipulating or masking the relevant attributes during different stages of training and evaluation. The proposed pipeline consists of three major phases:

#### 2.1.1 Phase 1. Full Shortcut Exclusion

We first prepare a controlled input—either training data that is carefully selected to exclude any shortcuts of sensitive attributes or other patterns whose influence we wish to study. The goal is to ensure that the model has no access to the attribute or pattern under investigation. The model is then trained using this controlled input, and the generated outputs are evaluated for further analysis.

For example, in the case of investigating whether data center-specific patterns influence the learning process, we exclude all data from a particular center during training and then evaluate the model’s performance on test data from that same center. This approach is also widely used in evaluating the generalizability of trained models, as it reveals how well a model can perform on completely unseen subsets associated with specific attributes. It is worth mentioning that, the test data would be balanced over both class types and sensitive groups. We consider the same test data over steps of our pipeline.

#### 2.1.2 Phase 2. Partial Shortcut Exclusion

In this phase, we exclude only a portion of the training data with target sensitive attribute or the shortcut pattern rather than completely removing it. The objective is to expose the model to limited cues, allowing us to observe whether partial exposure how affects the model’s behavior.

Building on the previous example, instead of excluding the entire data from a particular center, we now exclude only a specific class from that center while keeping other classes from the same center in the training set. During testing, we evaluate the excluded class from that center to observe whether it gets misclassified as one of the included classes belonging to target sensitive attribute.

#### 2.1.3 Phase 3. Deceptive Signal Measurement

In this phase, we aim to quantify the behavioral shift resulting from the evaluations performed in Phase 1 and Phase 2. The main goal is to compare how the model’s responses or predictions change when exposed to different levels of access to the sensitive attribute or shortcut pattern.

The method of comparison may vary depending on the specific application. For example, when evaluating how cancer-related features are influenced by data center-specific patterns in medical imaging, we may use classification accuracy shifts, confusion matrix disparities, or representation analysis. The next section presents a case study and the corresponding evaluation procedures used to implement and assess how the models trained to learn cancerous feature are reliable and make decisions based on truly target patterns.

### 2.2 Formalization

Let 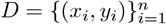 be a dataset containing samples *x*_*i*_ with class labels *y*_*i*_. Let *S*_*k*_ denote a sensitive subgroup (e.g., data from a specific data center), and let *C*_*i*_ represent a particular class label of interest.

#### 2.2.1 Phase 1. Full Exclusion

In this phase, we train a model 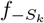 using all data in *D* except those belonging to subgroup *S*_*k*_, i.e. *D \ S*_*k*_.

We then evaluate 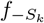 on 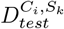, which is the subset of test data where the samples belong to class *C*_*i*_ and originate from subgroup *S*_*k*_. This setup allows us to measure how well the model generalizes to *S*_*k*_ when it has not seen any data from that subgroup during training. Then, we calculate 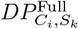 as the proportion of test samples from class *C*_*i*_ in subgroup *S*_*k*_ that are misclassified by the model 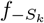.

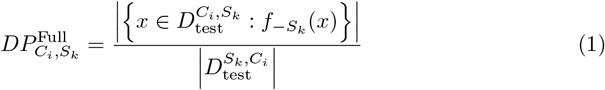

The expression of the numerator counts how many test samples of class *C*_*i*_ and subgroup *S*_*k*_ were misclassified by the model 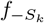, which was trained without seeing any samples from *S*_*k*_. The denominator is the total number of test samples from subgroup *S*_*k*_ and belonging to *C*_*i*_. This ratio quantifies the model’s error rate for a specific class–subgroup pair in a completely unseen subgroup scenario.

#### 2.2.2 Phase 2. Partial Exclusion

In second phase, we train model 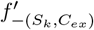 by excluding only class *C*_*ex*_ from *S*_*k*_ and including other classes belonging to *S*_*k*_ in the training set. Then, we test the model again on using the same test set, for each class into test test, to calculate the misclassified test samples by the model 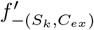.

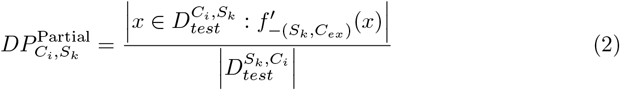

This equation measures how task-irrelevant features affect model performance by examining whether including partial classes causes the model to be deceived by irrelevant features embedded in those samples.

#### 2.2.3 Phase 3. Deceptive Signal Measurement

In this phase, we quantify the degree to which a model relies on shortcut patterns by comparing its behavior under two training conditions: full shortcut exclusion (as in Phase I) and partial shortcut exclusion (as in Phase II). The key idea is to measure how much the model’s predictions shift when it is trained with limited class access to a sensitive attribute, compared to when it has no access to that attribute across any class. In fact, in the second step, we use the included class types from *S*_*k*_ within training data to see weather model deceives in classification of test samples toward *S*_*k*_ specific features after including partial class of that test center. To capture this behavioral change, we define the Deceptive Disparity Shift 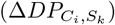 as the difference between the misclassification rate under partial and full exclusion for each class within the sensitive subgroup *S*_*k*_. Here, our objective is to analyze how the disparity shift of the excluded class within the training process is compared to the included classes of *S*_*k*_.

This is calculated as:

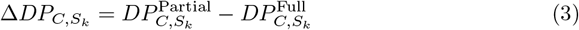

If the number of misclassified samples in the partial exclusion setting is greater than in the full exclusion setting, this suggests something important. In partial exclusion, the model has seen more data from the sensitive group *S*_*k*_, since only one class from *S*_*k*_ was left out during training. In contrast, in full exclusion, the model has seen no data at all from *S*_*k*_. So, if the model performs worse in the partial case, even with more related training data, it means the model is likely using patterns (or shortcuts) embedded in *S*_*k*_ samples from the seen class to make predictions about the unseen class in the same group instead of task relevant features. This behavior indicates that the model has learned to rely on group-specific features that are not truly relevant to the task.

To aggregate the calculated values across classes and derive the bias metric, we first compute the average disparity shift for the included classes within *S*_*k*_, denoted as 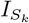, and treat this value as the baseline. This average is defined as:

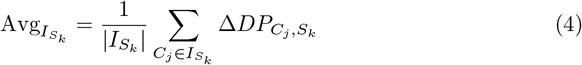

Then, the deceptive signal measure over *S*_*k*_ is then obtained by subtracting this baseline from the disparity shift of the excluded class within this sensitive group.

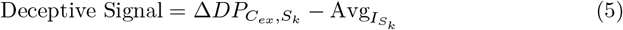

To interpret the bias metric meaningfully, it is useful to examine its behavior under extreme scenarios. Such scenarios reveal the theoretical limits of the metric and establish its lower and upper boundaries. We illustrate these limits through two contrasting cases: the *best case*, representing an unbiased model, and the *worst positive case*, reflecting strong reliance on shortcut patterns associated with the sensitive subgroup *S*_*k*_.

In the given equations, both *DP* ^Partial^ and *DP* ^Full^ are proportions of misclassified samples, and therefore take values in the range [0, 1]. This means that each 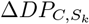 lies within [ −1, 1]. Because each 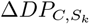 can only take values between −1 and 1, the average of these values across the included classes must also fall within the same range. Therefore, 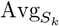 is bounded by −1 and 1.

With these bounds in place, the Deceptive Signal Measurement reaches its maximum value when the disparity shift for the excluded class is as large as possible 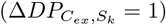 and the average disparity shift across included classes is as small as possible 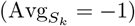. In this case:

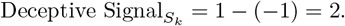

Similarly, the minimum value occurs when the disparity shift for the excluded class is at its lowest 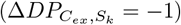 and the average disparity shift across included classes is at its highest 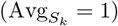, giving:

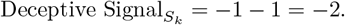

Thus, in general, the bias metric is bounded by:

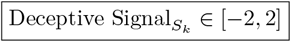

This range is universal and applies to all datasets and scenarios, regardless of the specific values of *DP* ^Partial^ and *DP* ^Full^.

## 3 Method Justification

This section justifies the Deceptive Bias Detection method using a probabilistic fairness. We aim to determine whether a model relies on sensitive subgroup-specific patterns (shortcuts) by comparing its behavior when such patterns are fully excluded versus when they are partially observed during training.

We define two training configurations:

- Full Exclusion: The model is trained without any samples from subgroup *S*_*k*_.

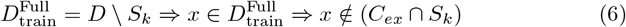
- Partial Exclusion: The model is trained with all samples from *S*_*k*_ except those from class *C*_*ex*_.

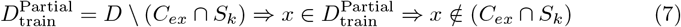

In both training datasets, 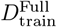 and 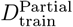 the model has not seen any samples from intersection (*C*_*ex*_, *S*_*k*_).

We now define two prediction distributions corresponding to the two exclusion settings:

- *P* (*f* (*x*) |∼ *S*_*k*_): The prediction behavior when the model is trained with no samples. from *S*_*k*_
- *P* (*f* (*x*) |∼ (*S*_*k*_ ∧ *C*_*ex*_)): The prediction behavior when the model is trained with samples from *S*_*k*_, excluding only those from *C*_*ex*_.

### Fairness Condition

If the model behaves fairly, meaning it does not rely on sensitive subgroup-specific patterns, then excluding all of *S*_*k*_ or just part of it should not change its behavior on *x* ∈ (*C*_*ex*_, *S*_*k*_). In this case, we should have:

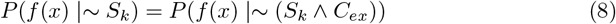

This equality is based on the assumption of conditional independence under fairness, which says that once we know the true class *C*_*ex*_, the model’s prediction should be independent of the sensitive attribute *S*_*k*_. Formally, this can be written as:

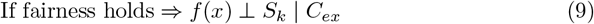

This means that, given the same class label, the subgroup from which the sample originates should not affect the model’s prediction. In a fair model, predictions are driven only by class-relevant features. Therefore, even when partial exposure to *S*_*k*_ is allowed (excluding only *C*_ex_), the model’s behavior on unseen samples from (*C*_*ex*_, *S*_*k*_) should remain the same as in the full exclusion case. However, if the model has learned shortcut features or latent signatures from the remaining data of *S*_*k*_ in the partial exclusion setting, then its behavior on will change, resulting in:

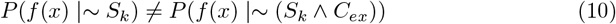

## 4 Case Study

To demonstrate the practical effectiveness of the Deceptive Bias Detection method, we conduct a case study using the TCGA histopathology dataset. The goal is to investigate whether the model relies on non-task-related patterns, specifically, data center-specific signatures, when performing cancer classification. The TCGA dataset [40] contains whole-slide histopathology images collected from multiple medical centers, each labeled with a cancer type. Prior research has shown that deep models trained on TCGA aimed at learning cancerous features can discriminate data site origin via cancerous features [11, 12]. To this end, we apply the methodology to this test case to investigate how reliable is the deep models’ learning process on these data samples. In our experiments, we fine-tune the EfficientNet architecture on various training subsets designed according to the Deceptive Bias Detection framework to assess whether the model utilizes center-based artifacts as shortcuts.

### 4.1 A Summary of Dataset and Deep Model

This study used the TCGA dataset, which includes 32,072 whole-slide images (WSIs) from 156 medical centers across 30 cancer types [41]. Each WSI was scanned at 20× magnification and divided into patches of 1000×1000 pixels. Low-quality or non-diagnostic regions were excluded following established protocols [41, 42]. For our analysis, we focussed on two types of cancer, lung adenocarcinoma (LUAD) and lung squamous cell carcinoma (LUSC), evenly sampled from three centers: Asterand, Prince Charles Hospital, and Roswell Park. To prepare the data, each patch was split into overlapping tiles of 224×224 pixels to match the input size of EfficientNet-B0, pre-trained on ImageNet. During training, we froze all layers except the last six for domain-specific fine-tuning. Additional layers included a MaxPooling2D, Flatten, and two dense layers (1024 and 256 units) with ReLU activation, dropout, and L2 regularization. A softmax layer was used for final cancer type classification, as presented at Fig. 1.

**Fig 1.**
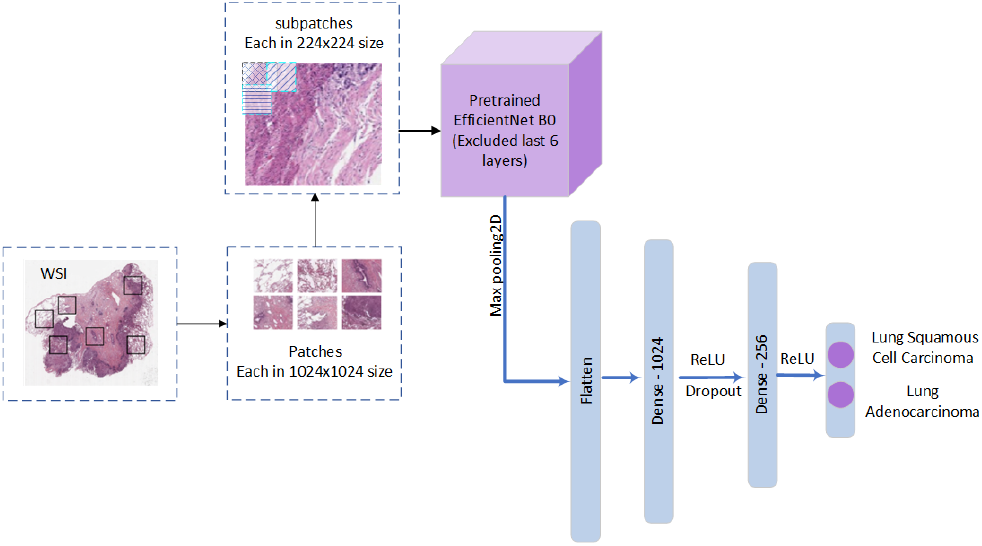
The figure shows how input patches are divided into smaller sub-patches to fit EfficientNet. Randomly chosen sub-patches are then passed through the pre-trained network, with only the last six layers kept trainable for adaptation.

### 4.2 Experimental Procedure on TCGA

First, we examine how origin-specific signatures in samples from ‘Asterand’ may influence the reliability and fairness of deep model training. For clarity, we present each step of the proposed method in detail.

### Phase 1. Full Exclusion

First, we exclude all data samples corresponding to *S*_*k*_ = *Asternad* from the training set, 𝒟_train_ = 𝒟 \ {*Asternad*}. Then, we use the remaining data to fine-tune the model. Of note, the samples of the other two date centers, which are evenly selected over both cancer types, are fixed during the whole training steps, the procedure is outlined in Fig 2.

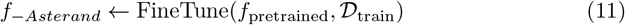

**Fig 2.**
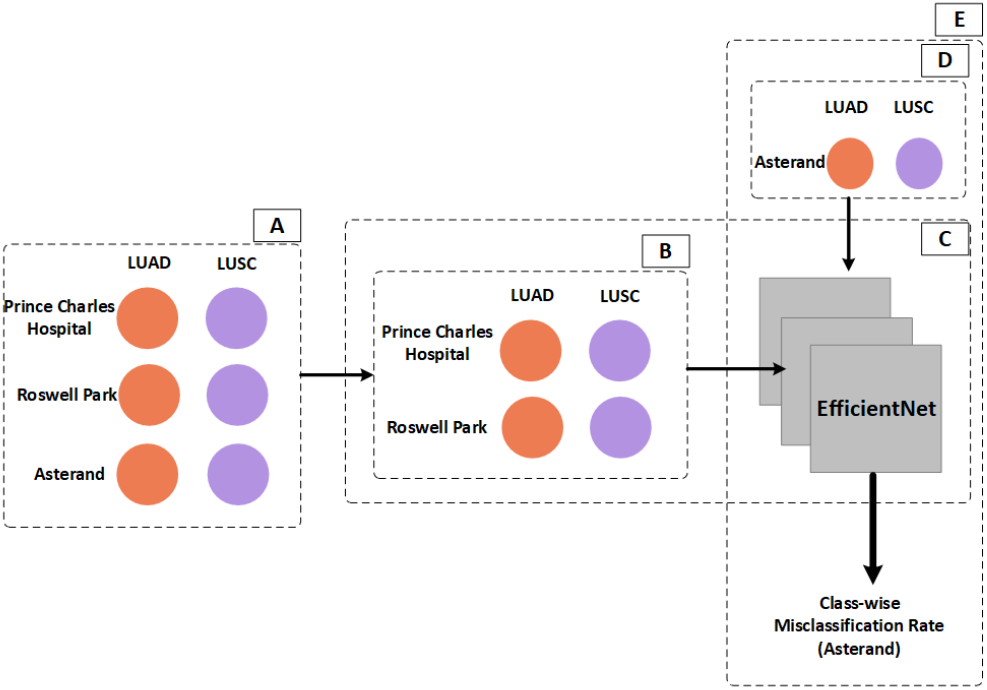
This figure represents the overview of the full Shortcut exclusion Scenario. A) The complete dataset includes LUAD and LUSC samples from three medical centers. B) The training set is constructed by excluding all Asterand samples. C) EfficientNet is trained only on Prince Charles and Roswell Park data. D) The model is then tested on the samples of the Asterand data center over both classes. E) The output reports the misclassification rate for each class, defined as the number of misclassified

Next, we apply the finetuned model on the data samples of both cancer types from ‘Asterand’ data center.

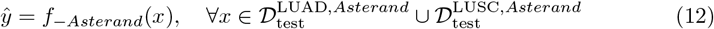

Then, for each cancer class (LUAD and LUSC ) included in 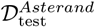, we calculate the proportion of samples that truly belong to LUAD or LUSC class, but categorized into LUSC or LUAD class out of total number of samples in 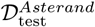. We consider the results achived in this step (completly removing data samples from Asterand data center) as the baseline to conduct further experiments. So, the misclassification rate or disparity proportion for LUAD and LUSC cancer types is calculated as shown in Eqs 13 and 14:

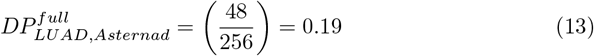

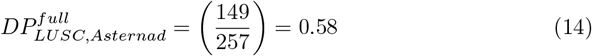

### Phase 2. Partial Exclusion

In this phase, referred to as partial data exclusion, outlined in Fig 3, we do not exclude all samples from the Asterand data center. Instead, we exclude only one cancer class from Asterand, while retaining the remaining classes from the same center in the training set. So, we decide to exclude data samples belonging to ‘Asterand’ with ‘LUAD’ cancer type, 𝒟_train_ = 𝒟 \ {*Asternad* ⋂ *LUAD*}. Next, we fine-tune the deep model using this modified dataset. In this setup, the model is exposed to only one cancer class from the ‘Asterand’ data center (LUSC), along with both cancer classes from the other two centers. For evaluation, we use the same test set as in the previous step, which includes samples from both cancer types originating from ‘Asterand’, to calculate the disparity proportion. Then, we can see how this partial exclusion affects the model’s performance over the same test samples of the LUAD class.

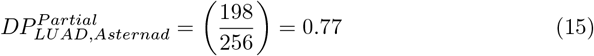

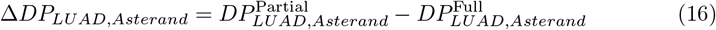

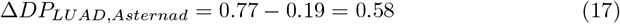

**Fig 3.**
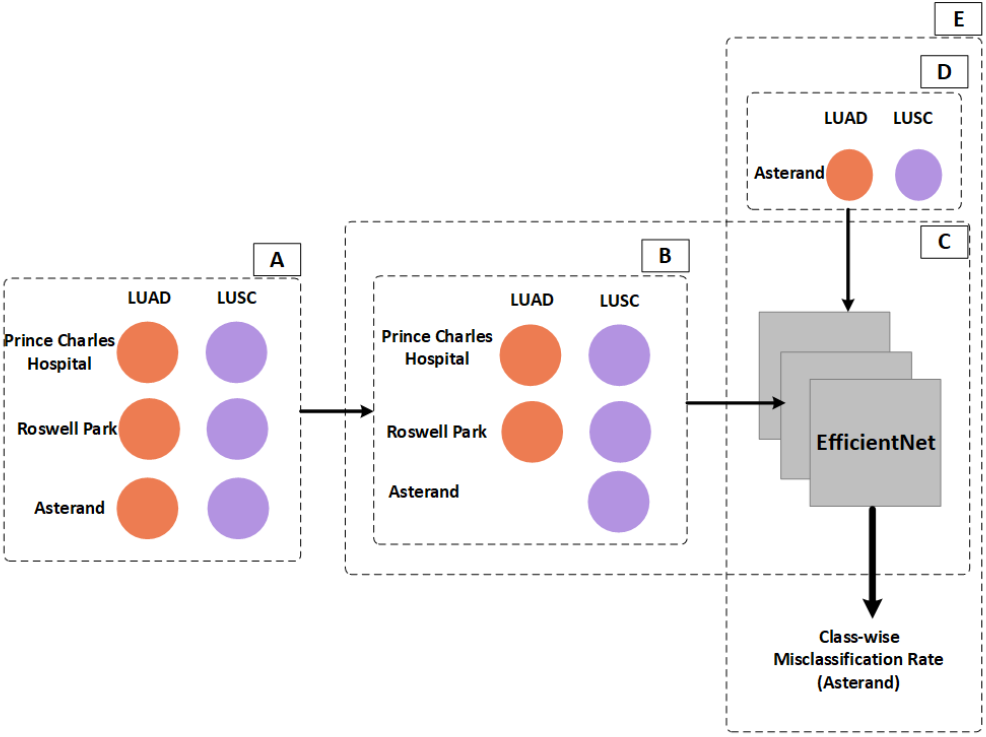
This figure represents the overview of the partial Shortcut exclusion Scenario. A) The complete dataset includes LUAD and LUSC samples from three medical centers. B) The training set is constructed by excluding only the LUAD class of the Asterand center. C) EfficientNet is trained only on Prince Charles and Roswell Park data. D) The model is then tested on the samples of the Asterand data center over both classes. E) The output reports the misclassification rate for each class, defined as the number of misclassified.

In this step, we observe that when the LUSC class from Asterand is included and the LUAD samples from this center are excluded, the number of LUAD samples misclassified as LUSC (the included class) increases significantly, from 48 to 198, as presented in Eq.s 13 and 15. This indicates that during training, the model captures center-specific signatures embedded in the included class, LUSC. As a result, when evaluating the model on unseen cancer types from the same center, it may rely on these spurious signatures for prediction, leading to incorrect classifications. This phenomenon mirrors the real-world example of a teacher designing exam questions. Modifying the training sample structure is similar to adjusting the tutorial questions used during instruction. The goal is to test whether students truly understand the underlying concepts or have merely memorized specific examples. In the next analysis, we evaluated the misclassified samples for the LUSC class after including its samples into training data. Only two test samples were misclassified, showing that including the LUSC class from the Asterand center significantly reduced misclassification compared to full exclusion.

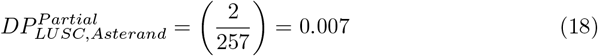

The corresponding Δ*DP* for this class is:

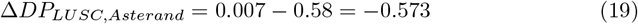

#### Phase 3. Deceptive Signal Measurement

We compare the disparity proportions under full and partial exclusion to quantify the reliance on center-specific shortcuts. By definition, Δ*DP* = *DP* ^Partial^ − *DP* ^Full^. For the LUAD class in the Asterand center, the misclassification rate increased sharply from 0.19 under full exclusion (Eq. 13) to 0.77 under partial exclusion (Eq. 15), yielding Δ*DP*_*LUAD,Asterand*_ = 0.58. This indicates that the model relied on center-specific signatures captured from the included LUSC class to classify LUAD samples, rather than learning cancer-relevant features.

In contrast, for the LUSC class, the misclassification rate dropped from 0.58 in the full exclusion setting (Eq. 14) to only 0.007 after partial inclusion (Eq. 18), giving Δ*DP*_*LUSC,Asterand*_ = −0.573. This shows that including the LUSC class during training reduced errors significantly, as the model had direct access to relevant class samples from Asterand.

Since we have only one included class, the baseline is set as 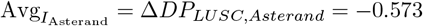. The Deceptive Signal for LUAD is then: 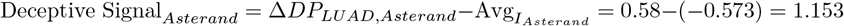.

This large positive value (within the theoretical range [ −2, 2]) highlights a strong deceptive effect: although including LUSC reduced misclassification for that class, it simultaneously biased predictions for the excluded LUAD class toward Asterand-specific patterns rather than true cancer features.

## 5 Discussion

This study introduces the Deceptive Signal metric as a practical way to check if deep learning models are truly learning the task or just memorizing hidden shortcuts. Unlike standard methods that only look at final accuracy scores, our approach tests how the model changes its behavior when we manipulate its training data. The core of this method lies in comparing two specific scenarios: “Full Exclusion,” where the model sees no data from a sensitive group (like a specific hospital), and “Partial Exclusion,” where it sees data from only some classes of that group. By observing the difference between these two stages, we can distinguish between a model that generalizes well and one that is “cheating.”

The logic behind this comparison is straightforward but powerful. In a normal learning process, giving a model more data, even if it is just from one class of a specific group, should help it understand the overall domain better. However, our method operates on the opposite hypothesis: if adding partial data actually makes the model perform worse on unseen samples from that same group, it is a clear sign of bias. This indicates that the model did not learn the actual features of the task (like cancer tissue) but instead learned a “shortcut,” such as the background noise unique to that hospital. This mirrors the “Math Teacher” analogy described earlier: if a student only memorizes the teacher’s specific given exercises in the trutorial sessions instead of learning the math concepts, they might pass a test on the memorized exercises rather than the underlying mathematical concepts, effectively quantifying the extent to which the model is “cheating” by exploiting these task-irrelevant shortcuts instead of learning the actual objective.

To make this measurement precise, we developed a quantitative metric that isolates this specific bias. By subtracting the average behavior of the classes the model did see from the behavior of the class it did not see, we ensure that we are measuring the shortcut reliance accurately. This calculation results in a score that is mathematically guaranteed to fall between -2 and 2. This standardized range is a significant advantage because it creates a universal “trust score” that can be used across different datasets. A score near +2 represents the worst-case scenario. This allows researchers and regulators to easily define safety limits, for example, deciding that any medical AI with a score above 0.5 is too risky to be used in real-world clinics.

## 6 Limitations

Despite the advantages of the Deceptive Signal metric, this study has specific limitations that must be acknowledged. First, the proposed method relies heavily on the availability of explicit labels for sensitive subgroups, such as the specific data center or hospital where an image was acquired. In many real-world datasets, this metadata may be anonymized, missing, or unreliable, which would prevent the construction of the necessary exclusion phases. Without these identifiable attributes, we cannot isolate the specific shortcuts the model might be learning.

Second, the methodology introduces a higher computational cost compared to traditional fairness metrics. Standard methods typically evaluate bias in a single pass after the model is fully trained. In contrast, our pipeline requires training the model multiple times under different conditions (Full Exclusion and Partial Exclusion) to calculate the behavioral shift. This requirement for multiple training runs increases the time and computing resources needed, which may be a constraint for large-scale models or resource-limited settings. Finally, while we demonstrated the effectiveness of this metric on histopathology images, the current study is limited to this specific domain, and further validation is needed to ensure its applicability to other types of data, such as text or natural images.

## 7 Future Work

Looking ahead, we see several promising directions to expand the utility of the Deceptive Signal metric. While this study focused on medical imaging, a critical next step is to adapt this framework for Large Language Models (LLMs). Like image classifiers, LLMs are prone to learning hidden shortcuts, often associating specific demographic cues or writing styles with certain answers rather than relying on factual content. By applying our exclusion–inclusion strategy to text data, we aim to verify if our metric can effectively quantify these “reasoning shortcuts” in natural language processing, thereby testing the universality of our approach.

Furthermore, we intend to move beyond simple detection and explore mitigation. Currently, our metric serves as a diagnostic tool to identify bias after training. In future research, we plan to integrate the Deceptive Signal calculation directly into the training process itself. By using this metric as a penalty term in the loss function, we could force the model to “unlearn” spurious correlations in real-time, effectively creating a “shortcut-averse” learning algorithm.

## Author contributions

F.K. is the corresponding author. M.M. and S.R. supervised the test case design. S.R. reviewed and edited the manuscript. F.K. made contributions to the design and implementation of test cases, results analysis, and manuscript drafting. All authors read and approved the final manuscript.

## Data availability

All the test cases were applied on the TCGA dataset publicity available at the following URL: “https://portal.gdc.cancer.gov“

